# Coronary bypass grafting in patients with three vessel coronary artery disease in the setting of ST-elevation myocardial infarction and Cardiogenic shock is associated with substantially lower mortality compared to undergoing percutaneous coronary intervention

**DOI:** 10.1101/2025.02.25.25322704

**Authors:** Mohammad Reza Movahed, Daniel McCoy, Mehrtash Hashemzadeh

**Author notes:** Correspondent: M Reza Movahed, MD, PhD, FACP, FACC, FSCAI, Clinical Professor of Medicine, University of Arizona Sarver Heart Center, 1501 North Campbell Avenue, Tucson, AZ 85724.

## Abstract

**Background:** The shock trial was the first small, randomized trial revealing revascularization of patients presenting with ST-elevation myocardial infarction (STEMI) with coronary bypass surgery (CABG) have a better survival rate than percutaneous coronary intervention (PCI) The goal of this study was to use the largest inpatients database, to evaluate inpatients mortality of STEMI patients with three-vessel disease and cardiogenic shock undergoing CABG vs PCI.

**Methode:** Using the Nationwide Inpatient Sample (NIS) database, and ICD-10 coding for cardiogenic shock, three-vessel CABG, and three-vessel PCI, we evaluate total inpatient mortality comparing three-vessel CABG vs three-vessel PCI in adults over age 18 years.

**Results:** A total of 3,565 STEMI patients with 3 vessel disease and cardiogenic shock underwent PCI vs.3,950 undergoing CABG. CABG in the setting of STEMI-related cardiogenic shock and three-vessel CAD is associated with much lower mortality compared to three-vessel PCI despite multivariate adjustment. Mortality was more than twice in patients undergoing PCI vs CABG (68.27% vs 31.73%, P<0.001, OR for CABG patients: 0.32, CI: 0.25-0.42, P<0.001). After adjusting baseline characteristics and comorbidities in multivariate analysis, CABG remained significantly associated with lower mortality (CABG OR 0.34, CI:0.26-0.44, p<0.001).

**Conclusion:** Our data confirms the finding of the Schock trial that three-vessel CABG is greatly superior to PCI in STEMI patients presenting with cardiogenic shock and three-vessel coronary artery disease.

**Condensed Abstract:** Using the Nationwide Inpatient Sample (NIS) database, and ICD-10 coding for cardiogenic shock, three-vessel CABG, and three-vessel PCI, we evaluate total inpatient mortality comparing three-vessel CABG vs three-vessel PCI in adults over age 18 years. A total of 3,565 STEMI patients with 3 vessel disease and cardiogenic shock underwent PCI vs.3,950 undergoing CABG. CABG in the setting of STEMI-related cardiogenic shock and three-vessel CAD is associated with much lower mortality compared to three-vessel PCI despite multivariate adjustment. After adjusting baseline characteristics and comorbidities in multivariate analysis, CABG remained significantly associated with lower mortality (CABG OR 0.34, CI:0.26-0.44, p<0.001).

## Introduction

Cardiogenic shock (CS) is defined as SBP□<□90 mm Hg for 30 minutes or inotrope use to maintain SBP□>□90 mmHg and increased filling pressures leading to systemic hypoperfusion and tissue hypoxia due to cardiac dysfunction [1, 2]. The most common etiology of CS is a myocardial infarction (MI) that causes cardiac myocyte necrosis and reduced ventricular function. CS occurs in 5–12% of patients following an MI and is the leading cause of mortality at approximately 40% at 30 days and 50% at 1 year [3,4]. With the current aging population in the United States, the incidence of CS is increasing, and patients are now more complex with more associated comorbidities [5].

The Should We Emergently Revascularize Occluded Coronaries for Cardiogenic Shock (SHOCK) trial demonstrated that there is a significant survival benefit with early revascularization when compared to medical management in the treatment of cardiogenic shock [6]. This led to a class 1 recommendation for emergency revascularization of patients with cardiogenic shock from the American College of Cardiology/American Heart Association [7]. In the SHOCK trial revascularization consisted of either percutaneous coronary intervention (PCI) in 55% or coronary artery bypass graft (CABG) in 38% of the patients [6].

Although both revascularization procedures are shown to be superior treatments compared to medical management, the relative merits and survival benefits of these revascularization methods may differ in patients with CS. Many patients with CS have complex three-vessel disease that may be treated better with CABG and PCI has a lower procedural success rate in patients with CS than in those without CS [8]. CABG may also provide the benefits through protection of ischemic myocardium and revascularization of noninfarct zones in patients with CS [8].

In this study, we aim to leverage data from a large National Inpatient Sample (NIS) database to evaluate and compare the inpatient survival benefits of PCI vs CABG in patients with three-vessel disease suffering from CS.

## Methods

### Data Source

This study is deemed institutional review board exempt as the NIS is a publicly available deidentified database. The NIS database includes weighted discharge information for about 35 million patients each year, 20% of all inpatient admissions to nonfederal hospitals in the United States.

### Study Population

Patient data was drawn for all patients over the age of 18 with a diagnosis of CS undergoing either three-vessel PCI or three-vessel CABG from the 2016-2020 database years. Both International Classification of Diseases, Tenth Revision, Clinical Modification (ICD-10-CM) and International Classification of Diseases, Tenth Revision, Procedure Coding System (ICD-10-PCS) codes were used to query the NIS database and develop the study cohort. The target population of patients having undergone PCI was identified using the ICD-10-PCS codes 02723(5-7)6, 02723(5-7)Z, 02723(D-G)Z, 02723(D-G)6, 02723T6, 02723TZ, 02723Z6, 02723ZZ, 02C23Z(6-7), 02C23ZZ. The target population of patients having undergone CABG was identified using the ICD-10-PCS codes 02120(8-9)3, 02120(8-9)8, 02120(8-9)9, 02120(8-9)C, 02120(8-9)F, 02120(8-9)W, 02120A3, 02120A8, 02120A9, 02120AC, 02120AF, 02120AW, 02120(J-K)3, 02120(J-K)8, 02120(J-K)9, 02120(J-K)C, 02120(J-K)F, 02120(J-K)W, 02120Z3, 02120Z8, 02120Z9, 02120ZC, 02120ZF, 0212344, 12123D4, 0212444, 02124(8-9)3, 02124(8-9)8, 02124(8-9)9, 02124(8-9)C, 02124(8-9)F, 02124(8-9)W, 02124A3, 02124A8, 02124A9, 02124AC, 02124AF, 02124AW, 02124D4, 02124(J-K)3, 02124(J-K)8, 02124(J-K)9, 02124(J-K)C, 02124(J-K)F, 02124(J-K)W, 02124Z3, 02124Z8, 02124Z9, 02124ZC, 02124ZF. These populations were further stratified using the ICD-10-CM codes I25.13 for three-vessel disease, 121.xx for MI, and R57.0 for shock. Cohort demographic data were calculated using age, sex, and race. After performing multivariate analysis on high-risk baseline features and characteristics, we added any characteristics that were significantly different between the 2 groups in the multivariate analysis.

### Study Outcomes

The patient outcome examined was inpatient total mortality. In multivariate analysis, we adjusted mortality for baseline characteristics and all high-risk features including age, race, diabetes, gender, chronic kidney disease, hyperlipidemia, chronic obstructive pulmonary disease, and smoking status

### Statistical Analysis

Patient demographic, clinical, and hospital characteristics are reported as means, with 95% confidence intervals for continuous variables and proportions, and 95% confidence intervals for categorical variables. Trend analysis over time was assessed using Chi-squared analysis for categorical outcomes and univariate linear regression for continuous variables. Multivariable logistic regression ascertained the odds of binary clinical outcomes relative to patient and hospital characteristics as well as the odds of clinical outcomes over time. All analyses were conducted following the implementation of population discharge weights. All P-values are 2-sided, and P < .05 was considered statistically significant. Data were analyzed using STATA 17 (StataCorp LLC, College Station, Texas).

## Results

A weighted total of 3,565 ST-elevation myocardial infarction (STEMI) patients with three-vessel disease and CS underwent PCI and 3,950 STEMI patients with three-vessel disease and CS underwent CABG from 2016-2020. The mean patient age for those who underwent PCI was 66.37 ± 11.82 years old; for those who underwent CABG mean age was 65.19 ± 10.37 years old.

White patients comprised the majority of the study cohort in both groups at 73.22% for the population of patients that underwent PCI and 69.97% for the population of patients that underwent CABG. Table 1 is showing baseline characteristics and comorbidities between PCI and CABG cohorts.

**Table 1.** Demographics and clinical characteristics of patients with ST-Elevation myocardial infarction associated cardiogenic shock undergoing three vessel coronary PCI (percutaneous coronary Intervention) vs CABG (Cornay bypass surgery).

| STEMI & Cardiogenic Shock patients with age above 18 |  |  |  |  |
| --- | --- | --- | --- | --- |
| 2016-2020 | PCI Three-Vessel | CABG Three-Vessel | P-value | Odds Ratio (C.I) |
| Total Population | 3,565 | 3,950 |  |  |
| Age |  |  | 0.04 |  |
| Mean±SD | 66.37±11.82 | 65.19±10.37 |  |  |
| Median(IQR) | 66(58-74) | 65.5(58-73) |  |  |
| LOS |  |  | <0.001 |  |
| Mean±SD | 9±10 | 13±11 |  |  |
| Median(IQR) | 6(2-11) | 11(7-17) |  |  |
| Total Charges \$ | | | 0.001 | |
| Mean±SD | 335939±462165 | 412111±375646 |  |  |
| Median(IQR) | 242771(152089-411343) | 306401(192083-507585.5) |  |  |
| Gender |  |  |  |  |
| Male | 69.52% | 30.48% |  | REF |
| Female | 77.59% | 22.41% | <0.001 | 0.66(0.53-0.83) |
| Race |  |  |  |  |
| White | 73.22% | 69.97% |  | REF |
| Black | 6.11% | 6.30% | 0.74 | 1.08(0.70-1.67) |
| Hispanic | 12.37% | 11.53% | 0.88 | 0.97(0.70-1.35) |
| Asian/Pac Isl | 3.64% | 5.63% | 0.07 | 1.62(0.96-2.72) |
| Native American | 0.44% | 0.67% | 0.52 | 1.61(0.38-6.77) |
| Others | 4.22% | 5.90% | 0.13 | 1.46(0.90-2.38) |
| Mortality | 68.27% | 31.73% | <0.001 | 0.32(0.25-0.42) |
| Diabetese | 45.22% | 54.78% | 0.12 | 1.17(0.96-1.43) |
| Smoking | 46.20% | 53.80% | 0.59 | 1.07(0.84-1.35) |
| CKD | 50.26% | 49.74% | 0.2 | 0.86(0.68-1.08) |
| COPD | 47.60% | 52.40% | 0.96 | 0.99(0.76-1.30) |
| Hyperlipidemia | 44.57% | 55.43% | 0.006 | 1.33(1.08-1.64) |
| Alcohol | 46.15% | 53.85% | 0.81 | 1.06(0.67-1.66) |

Univariate analysis showed that mortality was much lower in the group that underwent CABG at 31.73% compared to 68.27% in the PCI group (odds ratios [OR] 0.32; 95% CI, 0.25-0.42; P < 0.001). The CABG group also had higher rates of most of the complicating factors including smoking, hyperlipidemia, COPD, diabetes, and alcohol use. The length of stay was longer in the CABG patient population (median 13 days; SD 11) than in the PCI patient population (median 9; SD 10). The overall price was higher in the CABG patient population (median 412,111 dollars; SD 375,646) than in the PCI patient population (median 335,939 dollars; SD 462,165, Table 2.

**Table 2.** Univariate mortality, length of stay (LOS), and total charge of patients with ST-Elevation myocardial infarction associated cardiogenic shock undergoing three vessel coronary PCI (percutaneous coronary Intervention) vs CABG (Cornay bypass surgery).

|  | PCI Three-Vessel | CABG Three-Vessel | P-value | Odds Ratio<br>(C.I.) |
| --- | --- | --- | --- | --- |
| LOS |  |  | <0.001 |  |
| Mean±SD | 9±10 | 13±11 |  |  |
| Median(IQR) | 6(2-11) | 11(7-17) |  |  |
| Total Charges<br>\$ | | | 0.001 | |
| Mean±SD | 335939±462165 | 412111±375646 |  |  |
| Median(IQR) | 242771(152089-411343) | 306401(192083-507585.5) |  |  |
| Mortality | 68.27% | 31.73% | <0.001 | 0.32(0.25-0.42) |

Furthermore, after adjusting CABG intervention with numerous comorbidities such as diabetes, smoking, chronic kidney disease, chronic obstructive pulmonary disease, hyperlipidemia, age, sex, and race, CABG intervention remained independently associated with markedly lower mortality. (OR 0.34; 95% CI, 0.26-0.44; P < 0.001).

## Discussion

Each year in the United States, approximately 40,000 to 50,000 patients experience CS following an MI, making it the leading cause of in-hospital mortality among these patients [11–14]. Given the high prevalence and poor outcomes associated with CS, determining the optimal revascularization strategy for these patients is critical. Although research on this topic remains limited, existing data suggest that CABG is a viable revascularization option, particularly for patients with multivessel disease complicated by CS. In the SHOCK trial, patients undergoing CABG often presented with more severe coronary disease, including higher rates of diabetes, three-vessel disease, and left main disease. Nevertheless, their survival rates at 30 days and 1 year were comparable to those of patients treated with PCI [6]. Similarly, findings from the SHOCK Trial Registry revealed that patients undergoing CABG had lower in-hospital mortality compared to those receiving PCI, particularly in cases of two- and three-vessel disease [9].

Despite these benefits, CABG remains underutilized in the acute setting [18]. Although over 70% of patients with MI complicated by CS present with multivessel coronary artery disease, less than 4% undergo emergent CABG [15]. This underutilization may stem from logistical challenges, such as the difficulty of performing emergency CABG promptly and the higher associated costs compared to PCI.

Advancements in surgical techniques and patient care since the SHOCK trial have further improved CABG outcomes. For instance, the trial reported the use of left internal mammary arterial grafts in only 15.2% of cases [6]. Greater utilization of arterial grafts, alongside advances in mechanical support, cardioplegia, anesthesia, and patient selection, would likely contribute to better outcomes today. A study by Davierwala et al. documented a decline in hospital mortality rates for CABG in CS patients over time: from 42.2% (2000–2004) to 26.8% (2010–2014) [19]. The overall hospital mortality of 33.7% (2000-2014) in this cohort was lower than rates reported for PCI in the SHOCK trial and other studies, which range from 36% to 49% for multivessel disease in CS [19–21].

Our analysis of over 7,500 patients reinforces the evidence favoring CABG over PCI for patients with MI complicated by CS and three-vessel coronary artery disease. CABG was associated with dramatically lower mortality than PCI (31.73% vs. 68.27%, P < 0.001), with an odds ratio (OR) of 0.32 (CI: 0.25–0.42, P < 0.001). Even after adjusting for baseline characteristics and comorbidities, CABG maintained its association with markedly improved survival (OR: 0.34, CI: 0.26–0.44, P < 0.001). This survival benefit may be attributed to higher lower rates of incomplete revascularization associated with CABG compared to PCI (25% vs. 56%, P < 0.001) [16].

The importance of complete revascularization in CS has been supported by studies such as the Manitoba CS registry, which demonstrated improved hospital survival in patients with more complete revascularization [17]. A small propensity score analysis also showed that while PCI-alone patients had mortality rates consistent with prior trials (40.9%), those treated with both PCI and CABG had a markedly lower mortality rate of 20.5% [10].

Given the high prevalence of multivessel disease in CS and the suboptimal outcomes associated with multivessel PCI, emergency CABG should be more frequently considered in this patient population [22]. The CULPRIT-SHOCK trial underscored the challenges of multivessel PCI, reporting a 30-day mortality rate of 50% for CS patients, far exceeding rates observed with CABG in available studies [19,22]. However, while current data strongly favor CABG, randomized controlled trials are ultimately needed to definitively determine the optimal revascularization strategy for patients with MI complicated by CS.

## Limitations

Due to its retrospective design, this study has several limitations. Patients undergoing CABG were selected based on routine clinical decision-making rather than randomization, which could result in a lower-risk cohort. Additionally, as the study relied on procedure and diagnosis codes, misclassification due to coding errors cannot be ruled out.

## Data Availability

NIS data is publicly available

## Notes

Conflict of interest: None

### Competing Interest Statement

The authors have declared no competing interest.

### Funding Statement

None

